# Prevalence and patterns of different types of non-cigarette tobacco use in England: a population study

**DOI:** 10.1101/2025.05.12.25327445

**Authors:** Sarah E. Jackson, Eve Taylor, Harry Tattan-Birch, Katherine East, Lion Shahab, Sharon Cox, Masuma Pervin Mishu, David Hammond, Jamie Brown

**Affiliations:** Department of Behavioural Science and Health, University College London, UK; Department of Primary Care and Public Health, Brighton and Sussex Medical School, UK; National Addiction Centre, Institute of Psychiatry, Psychology & Neuroscience, Kings College London, UK; Behavioural Research UK, Edinburgh, UK; Department of Epidemiology and Public Health, University College London, UK; School of Public Health Sciences, University of Waterloo, Canada

**Keywords:** non-cigarette tobacco, cigars, cigarillos, chewing tobacco, waterpipe, heated tobacco

## Abstract

**Introduction:** Non-cigarette tobacco smoking increased in England between 2013 and 2023, but little is known about the prevalence of use of specific types of non-cigarette tobacco, including smokeless products, or whether the increasing trend has continued. This study examined recent trends in exclusive non-cigarette tobacco smoking and estimated the prevalence of different smoked and smokeless product use by sociodemographic characteristics, vaping and cigarette smoking status.

**Methods:** Data came from the Smoking Toolkit Study, a nationally representative monthly cross-sectional survey of adults (≥18y) in England (*n*=94,918; April-2020 to February-2025). We used logistic regression to estimate trends in exclusive non-cigarette tobacco smoking. We further examined the prevalence and correlates of (non-exclusive) use of eight specific smoked and smokeless non-cigarette tobacco products among 8,129 participants surveyed October-2024 to February-2025.

**Results:** The prevalence of exclusive non-cigarette tobacco smoking increased from 1.3% [1.1-1.6%] in April-2020 to 2.0% [1.8-2.1%] in August-2022, before declining to 1.2% [1.1-1.5%] by February-2025. Between October-2024 and February-2025, 3.7% [3.2-4.1%] of adults reported any non-cigarette tobacco use (∼1.7 million people). Cigars (1.2%) and waterpipes (0.9%) were the most used products. Use patterns varied: cigars, cigarillos, and pipes were more common among men and those who smoked cigarettes, while waterpipes were more common among younger adults, minority ethnic groups, and people who vaped. Smokeless tobacco use was more common among those who smoked (any form of) tobacco.

**Conclusions:** In England, exclusive non-cigarette tobacco smoking has fluctuated in recent years. Patterns of smoked and smokeless tobacco use vary across products and sociodemographic groups.

## Introduction

Tobacco use remains a leading cause of cancer and premature death.^1^ While cigarette smoking is the most common form of tobacco used globally,^1^ a wide range of non-cigarette tobacco products are also used, with marked regional variation – for example, snus (oral tobacco pouches) in Scandinavia^2^ and diverse oral tobacco products in parts of South Asia.^3^ In England, cigarettes smoking remains most prevalent^3^ and non-tobacco nicotine products like e-cigarettes and nicotine pouches are also widely available and popular.^4,5^ However, recent data suggest a notable rise in the use of non-cigarette smoked tobacco products (e.g., cigars, cigarillos, waterpipes [shisha], and pipes), especially since 2020.^3^ Additionally, some people use smokeless tobacco (e.g., snuff and different forms of chewing tobacco like betel quid [paan] with tobacco leaf or tobacco leaf mixture [zarda], dry tobacco powder [gul, mishri], khaini, gutka, etc.), snus (which cannot legally be sold in the UK), or heated tobacco products – and some use both smoked and smokeless forms. The health risks of non-cigarette tobacco use vary considerably from product to product – with smoked products generally more harmful – but all expose users to greater risk compared with using no tobacco products at all.^6–10^ Understanding the current prevalence of different forms of non-cigarette tobacco use, and how this differs across sociodemographic groups and regions of England, is important for monitoring trends and developing targeted interventions to reduce use where needed.

Culture and ethnicity are related to the use of some non-cigarette tobacco products. For example, waterpipe tobacco is predominantly used among Arab/Middle Eastern communities in the UK and is used to socialise and express cultural identity.^11–13^ Smokeless tobacco products, such as chewing tobacco, are primarily used by people from South Asian communities (predominantly among Indian, Pakistani, and Bangladeshi communities)^14^ with a range of different products used among different groups.^15^ Smokeless tobacco use is an important driver of inequality, causing higher rates of oral and pharyngeal cancers among South Asian ethnic groups compared with the general population in England.^16^ However, evidence suggests that ethnic minority groups and those aged 18-34 are more likely to underestimate the potential harms of non-cigarette tobacco products.^17^

Previous studies have reported descriptive data on non-cigarette tobacco use in England. For example, the International Tobacco Control (ITC) Survey found that among adults in 2020, the most commonly used smoked tobacco products (after cigarettes) were cigars, closely followed by waterpipes (shisha, hookah), pipes, and cigarillos.^18^ Among 16-19-year-olds in England, in 2023, cigarillos and waterpipe were the most popular tobacco products (after cigarettes), followed by cigars, smokeless tobacco, heated tobacco, and bidis.^19^ We recently published data from the Smoking Toolkit Study showing a substantial increase in the exclusive use of any smoked non-cigarette tobacco products in England between 2020 and 2023, particularly among younger adults (e.g., from 0.2% to 3.2% among 18-year-olds).^3^ Similar patterns have been observed in other datasets (e.g., the ITC Youth Survey), which show that, among youth in England, use of cigarillos has doubled since 2017, from 2.1% to 4.1% in 2023.^19^ HM Revenue and Customs figures also show increases in sales of cigars in the UK in recent years, from £73 million in 2019 to £165 million in 2024.^20^ In the context of this changing landscape, more up-to-date information is needed on current patterns of use, considering each different type of smoked and smokeless products.

This study aimed to provide an update on recent trends in exclusive non-cigarette tobacco smoking in England. It also aimed to estimate the current prevalence of any (i.e., non-exclusive) use of eight different smoked and smokeless non-cigarette tobacco products and how prevalence varies by sociodemographic characteristics, vaping status, and cigarette smoking status.

## Methods

### Pre-registration

The study protocol and analysis plan were pre-registered on Open Science Framework (https://osf.io/7vhk5/).

### Design

Data were drawn from the Smoking Toolkit Study, an ongoing monthly cross-sectional survey of adults (≥16 years) in England.^21,22^ The study began in 2006 and is ongoing. Data were originally collected via face-to-face interviews, but data collection switched to telephone interviews from April 2020 onwards. The study uses a hybrid of random probability and simple quota sampling to select a new sample of approximately 1,700 adults in England each month. Comparisons with other national surveys and sales data indicate the survey achieves nationally representative estimates of key variables such as sociodemographic characteristics, smoking prevalence, and cigarette consumption.^21,23^

The survey asks participants about their smoking status, which includes a response option that indicates exclusive non-cigarette tobacco smoking (i.e., that the participant does not use cigarettes but uses some other form of smoked tobacco; see *Measures*). Analyses of trends in the prevalence of exclusive non-cigarette tobacco smoking used data from those surveyed between April 2020 (the first wave to collect data via telephone, for consistency across the time series) and February 2025 (the most recent data available at the time of analysis).

Recently, we expanded the survey to include additional questions on the use of different smoked and smokeless non-cigarette tobacco products. Analyses of the current prevalence of specific forms of non-cigarette tobacco use used data from those surveyed between October 2024 and February 2025 (the only waves to include these questions).

For all analyses, we restricted the sample to participants aged ≥18 years (the legal age of sale of tobacco products in England).

### Measures

Full details of the measures are provided in the study protocol (https://osf.io/7vhk5/).

### Smoking status

Smoking status was assessed by asking participants which of the following best applied to them: (a) I smoke cigarettes (including hand-rolled) every day; (b) I smoke cigarettes (including hand-rolled), but not every day; (c) I do not smoke cigarettes at all, but I do smoke tobacco of some kind (e.g. pipe, cigar or shisha); (d) I have stopped smoking completely in the last year; (e) I stopped smoking completely more than a year ago; (f) I have never been a smoker (i.e. smoked for a year or more). Those who responded c were considered to exclusively smoke non-cigarette tobacco. For some analyses, we categorised responses as current tobacco smoking (a-c), former smoking (d-e) and never regular smoking (f).

### Use of different non-cigarette tobacco products

We assessed the use of a range of smoked and smokeless tobacco products. Smoked products included pipes, cigars, cigarillos, waterpipes (commonly referred to as shisha, narghile or hookah), and bidis. Smokeless products included snus; heated tobacco products; and other smokeless tobacco (chewing tobacco, paan, gutka, snuff, and dip). We did not analyse the use of tobacco-free nicotine products, such as e-cigarettes or nicotine pouches.

Use of heated tobacco products is assessed routinely in the Smoking Toolkit Study survey within several questions that ask about use of a range of nicotine products. Those who currently smoked were asked ‘Do you regularly use any of the following in situations when you are not allowed to smoke?’ and those who reported cutting down ‘Which, if any, of the following are you currently using to help you cut down the amount you smoke?’; those who had smoked in the past year were asked ‘Can I check, are you using any of the following either to help you stop smoking, to help you cut down or for any other reason at all?’; and those who had not smoked in the past year were asked ‘Can I check, are you using any of the following?’. Those who reported using a ‘heat-not-burn cigarette (e.g. iQOS with HEETS, heatsticks)’ in response to any of these questions were considered to use heated tobacco products.

Use of other products was assessed via new questions included in the survey between October 2024 and February 2025. Those who did not report exclusive non-cigarette smoking (i.e., responses a-b or d-f to the item assessing smoking status) were asked: ‘Can I check, are you using any of the following?’ Participants were asked to select all that applied from the following options: ‘pipe’, ‘cigar’, ‘cigarillo’, ‘shisha/waterpipe/hookah’, ‘bidis’, ‘chewing tobacco/paan/gutka/snuff/dip’, ‘snus’, ‘none of these’. For those who reported exclusive non-cigarette smoking (i.e., response c), these response options were split across two questions. The first asked about the type(s) of combustible products they used: ‘Can I just check, which of the following tobacco products do you smoke?’, with response options: ‘pipe’, ‘cigar’, ‘cigarillo’, ‘shisha/waterpipe/hookah’, ‘bidis’, ‘other’. The second asked whether they used any smokeless tobacco products: ‘Can I check, are you using any of the following?’, with response options: ‘chewing tobacco/paan/gutka/snuff/dip’, ‘snus’, ‘none of these’, ‘other’. For each of these questions, participants were asked to select all that applied.

### Sociodemographic characteristics and vaping status

Sociodemographic characteristics included age, gender, socioeconomic position (indexed by occupational social grade), ethnicity, and region in England.

Age was analysed as a continuous variable for trend analyses, modelled using restricted cubic splines (see statistical analysis section). We also provided descriptive data by age group (18-29/30-64/≥65).

Gender was self-reported as man, woman, or in another way; the latter group were excluded from regression analyses by gender due to low numbers (mean [SD] *n* per wave = 12 [5]).

Occupational social grade was categorised as ABC1 (managerial, professional, and upper supervisory occupations) or C2DE (manual routine, semi-routine, lower supervisory, state pension, and long-term unemployed), using the National Readership Survey classification system.^24^

Ethnicity was categorised according to the UK Census 2021 classification^25^ as White (White British, White Irish, White Gypsy/Traveller, White Other), Black (Black African, Black Caribbean, Black Other), Asian (Asian Indian, Asian Pakistani, Asian Bangladeshi, Asian Chinese, Asian Other), Mixed or Multiple ethnicities (Mixed White/Black Caribbean, Mixed White/Black African, Mixed White/Asian, Mixed Other), or Other (Arab, Other). For regression analyses, we collapsed these categories to white or minority ethnic group, due to low numbers in minority groups. Data on ethnicity were not collected between April-August 2020, so analyses by ethnicity excluded these waves.

Region was categorised as North (North West, North East, Yorkshire and the Humber), Midlands (West Midlands, East Midlands, East of England), or South (South West, South East, London). We also reported descriptive data on the prevalence of product use separately within each of the nine regions.

Vaping status was categorised as current vs. other, assessed with the same series of questions used to assess heated tobacco use. Those who reported using an e-cigarette in response to any of these questions were considered to currently vape.

### Statistical analysis

Data were analysed using R v.4.4.1. The Smoking Toolkit Study uses raking to weight the sample to match the population in England. This profile is determined each month by combining data from the UK Census, the Office for National Statistics mid-year estimates, and the annual National Readership Survey.^21^ The following analyses used weighted data. We excluded participants who did not report their smoking status; missing data on other variables (**Table S1**) were excluded on a per-analysis basis.

### Trends in exclusive non-cigarette tobacco smoking, 2020-2025

Consistent with our previous work,^3^ we used binary logistic regression to model trends in exclusive non-cigarette tobacco smoking (coded 1 for those who responded c to the question assessing smoking status and 0 for all other responses) between April 2020 and February 2025, overall and by sociodemographic characteristics.

For the overall model, the independent variable was time (survey wave), modelled using restricted cubic splines, to allow for flexible and non-linear changes over time while avoiding categorisation. We compared models with time analysed using restricted cubic splines with three, four, and five knots (sufficient to accurately model trends across years without overfitting) using the Akaike Information Criterion (AIC). Three knots provided the best fitting model (AIC=16107 vs. 16110 and 16112 for four and five knots, respectively).

For models testing moderation of trends by age, gender, occupational social grade, ethnicity, region, and vaping status, we included the interaction between the moderator of interest and time – thus allowing for time trends to differ across subgroups. Each of the interactions was tested in a separate model, without adjustment for other variables. Age was modelled using restricted cubic splines with three knots (placed at the 5, 50, and 95% percentiles), to allow for a non-linear relationship between age and exclusive non-cigarette tobacco smoking.

We used predicted estimates from these models to plot the prevalence of exclusive non-cigarette tobacco smoking over the study period, among all adults and within each subgroup of interest. We also reported modelled estimates of prevalence (with 95% confidence intervals [CI]) in April 2020 and February 2025 (the first and last months of the period), overall and within each subgroup.

### Prevalence of any current use of different non-cigarette tobacco products, 2024/25

Using data collected between October 2024 and February 2025 on specific products used, we reported the prevalence (aggregated across survey waves; with 95% CI) of any (i.e., non-exclusive) current use of: (i) any non-cigarette tobacco product, (ii) any smoked non-cigarette tobacco product, and (iii) any smokeless non-cigarette tobacco product. We then reported the specific prevalence of use of pipes, cigars, cigarillos, waterpipes [shisha/waterpipe/hookah], bidis, snus, heated tobacco products, and other smokeless tobacco [chewing tobacco/paan/gutka/snuff/dip].

We reported estimates for all adults and stratified by sociodemographic characteristics, vaping status, and current cigarette smoking (yes/no; for analyses of smoked non-cigarette product use) or tobacco smoking status (current/former/never; for analyses of smokeless non-cigarette product use). We also reported the proportion (with 95% CI) of adults who dual used (i) cigarettes and non-cigarette smoked tobacco and (ii) smoked (including cigarettes) and smokeless tobacco products.

We then used binary logistic regression models to explore unadjusted and independent (adjusted for other potentially influencing factors) associations with product use. For each product category, we constructed models with use of the product as the dependent variable and age, gender, occupational social grade, ethnicity, region, vaping status, and current cigarette smoking (analyses of smoked products)/smoking status (analyses of smokeless products) as independent variables. Unadjusted models tested associations of product use with each independent variable separately. Adjusted models included all independent variables together, to determine the association of each variable after accounting for the others. We focus our discussion on independent associations, given word constraints.

In an unplanned (i.e., not pre-registered) analysis, we explored unadjusted differences in use of snus between participants who did and did not report current use of tobacco-free nicotine pouches (assessed using the same questions used to capture use of heated tobacco products).

## Results

A total of 95,589 adults (≥18y) in England responded to the survey between April 2020 and February 2025. We excluded 671 (0.7%) who did not report their smoking status, leaving a final sample of 94,918 participants for analyses of trends in exclusive non-cigarette tobacco smoking (non-exclusive use was not assessed before October 2024). Of these, 8,129 were surveyed between October 2024 and February 2025 and formed the sample for descriptive analyses of the prevalence of use of eight different (smoked and smokeless) non-cigarette tobacco products.

Characteristics of both samples are summarised in **Table S1**.

### Trends in exclusive non-cigarette tobacco smoking

**Figure 1** shows modelled time trends in the weighted proportion of adults reporting exclusive non-cigarette tobacco smoking between 2020 and 2025. **Table 1** shows modelled weighted prevalence estimates in the first and last months of the study period.

**Figure 1.**
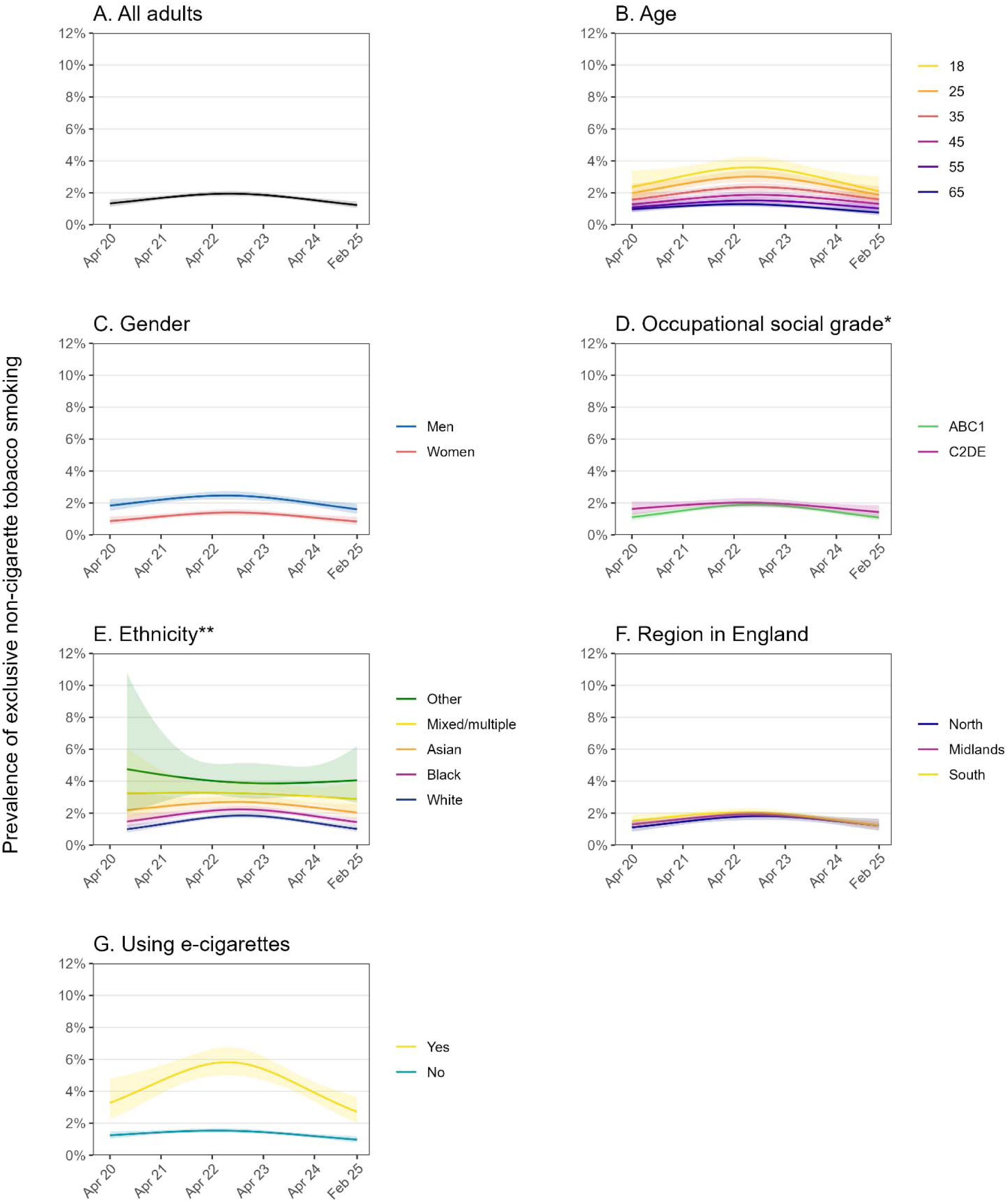
Trends in exclusive non-cigarette tobacco smoking, overall and within subgroups of adults in England, April 2020 to February 2025. Panels show trends (A) overall and by (B) age, (C) gender, (D) occupational social grade, (E) ethnicity, (F) region in England, and (G) vaping status. Lines represent modelled weighted prevalence by monthly survey wave, modelled non-linearly using restricted cubic splines (three knots). Shaded bands represent 95% confidence intervals. * ABC1 = more advantaged; C2DE = less advantaged ** Ethnicity was not assessed in April – August 2020; the model therefore used data from September 2020 – February 2025.

**Table 1.** Modelled estimates of the prevalence of exclusive non-cigarette tobacco smoking among adults in England, April 2020 to February 2025 (*n*=94,918)

| Characteristic | Exclusive non-cigarette tobacco smoking,<br>row % [95% CI]<sup>1</sup> |  |
| --- | --- | --- |
|  | April 2020 | February 2025 |
| All adults | 1.3 [1.1–1.6] | 1.2 [1.1–1.5] |
| Age (years)<sup>2</sup> |  |  |
| 18 | 2.4 [1.6–3.4] | 2.1 [1.5–3.0] |
| 25 | 2.0 [1.5–2.6] | 1.9 [1.5–2.4] |
| 35 | 1.6 [1.3–1.9] | 1.6 [1.3–1.9] |
| 45 | 1.3 [1.0–1.6] | 1.3 [1.0–1.7] |
| 55 | 1.1 [0.8–1.4] | 1.0 [0.8–1.3] |
| 65 | 1.0 [0.8–1.2] | 0.8 [0.6–1.0] |
| Age group (years)<sup>3</sup> |  |  |
| 18–29 | 2.1 [1.5–2.8] | 1.9 [1.5–2.5] |
| 30–64 | 1.3 [1.0–1.6] | 1.3 [1.0–1.6] |
| ≥65 | 0.9 [0.6–1.4] | 0.5 [0.3–0.9] |
| Gender<sup>4</sup> |  |  |
| Men | 1.8 [1.5–2.2] | 1.6 [1.3–2.0] |
| Women | 0.9 [0.6–1.2] | 0.8 [0.6–1.1] |
| Occupational social grade |  |  |
| ABC1 (more advantaged) | 1.1 [0.9–1.4] | 1.1 [0.9–1.3] |
| C2DE (less advantaged) | 1.6 [1.3–2.1] | 1.4 [1.1–1.9] |
| Ethnicity<sup>5</sup> |  |  |
| White | 1.0 [0.8–1.3] | 1.0 [0.8–1.2] |
| Black | 1.5 [1.1–1.9] | 1.4 [1.2–1.7] |
| Asian | 2.2 [1.4–3.4] | 2.0 [1.6–2.6] |
| Mixed/multiple | 3.2 [1.7–6.1] | 2.9 [2.1–4.0] |
| Other | 4.8 [2.0–10.8] | 4.1 [2.6–6.2] |
| Region in England |  |  |
| North | 1.1 [0.8–1.5] | 1.2 [0.9–1.7] |
| Midlands | 1.3 [1.1–1.5] | 1.2 [1.0–1.5] |
| South | 1.5 [1.2–1.9] | 1.2 [1.0–1.6] |
| Vaping status |  |  |
| Current vaping | 3.3 [2.2–4.8] | 2.7 [2.0–3.6] |
| Not | 1.2 [1.0–1.5] | 1.0 [0.8–1.2] |

CI, confidence interval.
<sup>1</sup> Weighted estimates of prevalence from logistic regression with survey month (April 2020 – February 2025) modelled non-linearly using restricted cubic splines. Note that data were modelled across this whole period; data reported here highlight the estimates in the first and last months. See **Figure 1** for estimates in each month. The outcome is non-cigarette smoking exclusive of cigarette smoking (i.e., excludes participants concurrently smoking cigarettes), rather than exclusive of smokeless tobacco use (i.e., does not exclude participants concurrently using smokeless tobacco products).
<sup>2</sup> Note that the model used to derive these estimates included data from participants of all ages, not only those who were aged exactly 18, 25, 35, 45, 55, or 65 years.
<sup>3</sup> Calculated as the mean of modelled estimates for the ages in each age group.
<sup>4</sup> Excludes 694 participants who described their gender in another way. Prevalence in this group aggregated across all survey months was 1.7% [1.6–1.8%].
<sup>5</sup> Ethnicity was not assessed in April – August 2020; the model therefore used data from September 2020 – February 2025.

The prevalence of exclusive non-cigarette tobacco smoking followed a non-linear trend (**Figure 1A**). It increased from 1.3% [1.1–1.6%] in April 2020 to a high of 2.0% [1.8–2.1%] in August 2022, then decreased to 1.2% [1.1–1.5%] by February 2025. As a result, there was little overall change from the start to the end of the period (**Table 1**). This did not appear to be driven by changes in cigarette smoking; a similar pattern was observed among adults who smoked tobacco and among adults in general (**Figure S1**).

There were some subgroup differences (**Figure 1B-G**). Across the period, the prevalence of exclusive non-cigarette tobacco smoking was consistently higher among participants who were younger, men, from minority ethnic groups (in particular, mixed/multiple and other ethnicities), and those who reported current vaping. The increase and subsequent decline in prevalence were greatest among those who were younger and among those who reported current vaping. There were also differences by ethnicity, with no notable change over time among those from mixed/multiple and other ethnicities. Time trends were similar by gender, occupational social grade, and region. Across all population subgroups, there was no notable difference in prevalence in February 2025 compared with April 2020 (**Table 1**).

### Prevalence of any current use of different non-cigarette tobacco products

**Table 2** shows the weighted prevalence of any (i.e., non-exclusive) current use of (i) any, (ii) any smoked, and (iii) any smokeless non-cigarette tobacco product, among participants surveyed between October 2024 and February 2025 (*n*=8,129), overall and within population subgroups. **Table 3** shows the weighted prevalence of the eight specific smoked and smokeless non-cigarette products over this period. **Tables S2-S5** provide a more detailed breakdown by ethnicity and region. **Tables 4**-**5** show independent associations between current use of each product and age, gender, occupational social grade, ethnicity, region, vaping status, and cigarette smoking (for associations with smoked product use) or tobacco smoking status (for associations with smokeless product use); unadjusted associations are summarised in **Tables S6-S7**.

**Table 2.** Prevalence of any (i.e., non-exclusive) current use of non-cigarette tobacco among adults in England, October 2024 – February 2025 (*n*=8,129; data aggregated across waves)

|  | Prevalence<sup>1</sup>, row % [95%CI] |  |  |
| --- | --- | --- | --- |
|  | Any non-cigarette tobacco product(s)<sup>2,3</sup> | Any smoked non-cigarette tobacco product(s)<sup>3,4</sup> | Any smokeless non-cigarette tobacco product(s)<sup>5</sup> |
| All adults | 3.7 [3.2–4.1] | 2.6 [2.2–3.0] | 1.2 [1.0–1.5] |
| Age (years) |  |  |  |
| 18-29 | 7.6 [6.1–9.1] | 4.0 [2.9–5.1] | 4.0 [2.9–5.1] |
| 30-64 | 3.2 [2.7–3.8] | 2.7 [2.2–3.2] | 0.7 [0.4–0.9] |
| ≥65 | 1.4 [0.8–1.9] | 1.1 [0.7–1.6] | 0.3 [0.0–0.5] |
| Gender |  |  |  |
| Men | 5.1 [4.3–5.8] | 3.7 [3.0–4.3] | 1.7 [1.2–2.1] |
| Women | 2.1 [1.6–2.6] | 1.4 [1.0–1.9] | 0.7 [0.4–1.0] |
| In another way | 10.3 [2.9–17.7] | 7.4 [1.0–13.7] | 2.9 [0.0–7.1] |
| Occupational social grade |  |  |  |
| ABC1 (more advantaged) | 3.6 [3.1–4.2] | 2.4 [2.0–2.8] | 1.4 [1.0–1.7] |
| C2DE (less advantaged) | 3.7 [2.9–4.5] | 2.8 [2.1–3.5] | 1.0 [0.6–1.5] |
| Ethnicity<sup>6</sup> |  |  |  |
| White | 3.3 [2.8–3.7] | 2.3 [1.9–2.7] | 1.1 [0.8–1.4] |
| Black | 6.4 [3.8–8.9] | 4.2 [2.2–6.2] | 2.3 [0.7–3.9] |
| Asian | 5.1 [3.0–7.2] | 4.4 [2.4–6.3] | 1.1 [0.1–2.1] |
| Mixed or multiple | 5.3 [2.4–8.3] | 3.5 [1.2–5.8] | 2.2 [0.1–4.4] |
| Other | 6.5 [1.9–11.1] | 4.2 [0.2–8.1] | 2.4 [0.0–4.8] |
| Region in England<sup>7</sup> |  |  |  |
| North | 3.9 [3.0–4.8] | 3.1 [2.3–4.0] | 1.1 [0.6–1.5] |
| Midlands | 3.1 [2.3–3.9] | 2.0 [1.4–2.6] | 1.2 [0.7–1.7] |
| South | 3.9 [3.2–4.6] | 2.6 [2.1–3.2] | 1.3 [0.9–1.7] |
| Vaping status |  |  |  |
| Current vaping | 10.0 [8.0–12.0] | 6.9 [5.2–8.6] | 3.8 [2.6–5.0] |
| Not | 2.7 [2.2–3.1] | 1.9 [1.6–2.3] | 0.8 [0.6–1.1] |
| Smoking status |  |  |  |
| Current smoking<sup>8</sup> | 16.8 [14.4–19.1] | 13.9 [11.8–16.1] | 3.6 [2.4–4.8] |
| Cigarette smoking | 9.4 [7.5–11.3] | 6.3 [4.7–7.9] | 3.9 [2.6–5.1] |
| Exclusive non-cigarette tobacco smoking | 100 [100–100] | 100 [100–100] | 0.6 [0.0–1.8] |
| Former smoking | 1.8 [1.2–2.4] | 0.4 [0.2–0.7] | 1.4 [0.9–2.0] |
| Never regularly smoked | 1.0 [0.7–1.3] | 0.6 [0.3–0.8] | 0.5 [0.3–0.7] |

<sup>1</sup> Prevalence of any smoked and any smokeless product use do not necessarily sum to the prevalence of any product use, because people can be dual users of smoked and smokeless products.
<sup>2</sup> Defined as current use of pipes, cigars, cigarillos, waterpipes, bidis, snus, heated tobacco, or other smokeless tobacco.
<sup>2</sup> Also includes participants who reported (separately) that they did not currently smoke cigarettes but smoked some other form of tobacco.
<sup>3</sup> Any current use of pipes, cigars, cigarillos, waterpipes, or bidis.
<sup>4</sup> Any current use of snus, heated tobacco, or other smokeless tobacco.
<sup>5</sup> Estimates by a more detailed ethnic breakdown are provided in **Table S2**.
<sup>6</sup> Estimates within each region in the North, Midlands, and South are provided in **Table S4**.
<sup>7</sup> Current smoking includes participants who reported smoking cigarettes or other tobacco products.
Note: Empty cells indicate where no participants in the subgroup reported use of the product.

**Table 3.** Prevalence of any (i.e., non-exclusive) current use of different forms of non-cigarette tobacco among adults in England, October 2024 – February 2025 (*n*=8,129; data aggregated across waves)

|  | Prevalence, row % [95%CI] |  |  |  |  |  |  |  |
| --- | --- | --- | --- | --- | --- | --- | --- | --- |
|  | Pipe | Cigar | Cigarillo | Waterpipe | Bidis | Snus | Heated tobacco | Other smokeless tobacco<sup>1</sup> |
| All adults | 0.4 [0.2–0.5] | 1.2 [0.9–1.4] | 0.6 [0.4–0.8] | 0.9 [0.7–1.2] | 0.1 [0.0–0.2] | 0.6 [0.5–0.8] | 0.3 [0.1–0.4] | 0.4 [0.2–0.6] |
| Age (years) |  |  |  |  |  |  |  |  |
| 18-29 | 0.4 [0.1–0.7] | 1.7 [0.9–2.4] | 0.9 [0.4–1.5] | 2.3 [1.4–3.1] | 0.3 [0.0–0.7] | 2.6 [1.8–3.5] | 0.5 [0.1–0.9] | 1.1 [0.5–1.8] |
| 30-64 | 0.4 [0.1–0.6] | 1.2 [0.9–1.5] | 0.6 [0.3–0.8] | 0.8 [0.5–1.1] | 0.0 [0.0–0.1] | 0.2 [0.1–0.3] | 0.3 [0.1–0.4] | 0.2 [0.1–0.4] |
| ≥65 | 0.3 [0.0–0.7] | 0.7 [0.3–1.0] | 0.3 [0.1–0.5] | 0.1 [0.0–0.2] | 0.1 [0.0–0.2] | 0.0 [0.0–0.1] | 0.1 [0.0–0.2] | 0.2 [0.0–0.4] |
| Gender |  |  |  |  |  |  |  |  |
| Men | 0.5 [0.2–0.7] | 2.1 [1.7–2.6] | 0.9 [0.5–1.2] | 1.2 [0.8–1.6] | 0.1 [0.0–0.2] | 1.0 [0.6–1.3] | 0.3 [0.1–0.4] | 0.5 [0.3–0.8] |
| Women | 0.1 [0.0–0.3] | 0.2 [0.0–0.3] | 0.2 [0.1–0.4] | 0.6 [0.4–0.8] | 0.1 [0.0–0.3] | 0.3 [0.1–0.4] | 0.2 [0.1–0.4] | 0.2 [0.0–0.3] |
| In another way | 2.9 [0.0–7.1] | 2.9 [0.0–7.1] | 2.9 [0.0–7.1] | 4.4 [0.0–9.4] | - | - | - | 2.9 [0.0–7.1] |
| Occupational social grade |  |  |  |  |  |  |  |  |
| ABC1 (more advantaged) | 0.3 [0.2–0.5] | 1.2 [0.9–1.5] | 0.5 [0.3–0.7] | 1.0 [0.7–1.2] | 0.1 [0.0–0.3] | 0.8 [0.6–1.1] | 0.3 [0.2–0.5] | 0.3 [0.1–0.5] |
| C2DE (less advantaged) | 0.4 [0.1–0.7] | 1.1 [0.7–1.5] | 0.7 [0.3–1.0] | 0.9 [0.5–1.3] | 0.1 [0.0–0.2] | 0.4 [0.1–0.7] | 0.2 [0.0–0.4] | 0.5 [0.2–0.8] |
| Ethnicity<sup>2</sup> |  |  |  |  |  |  |  |  |
| White | 0.4 [0.2–0.6] | 1 [0.8–1.3] | 0.5 [0.3–0.7] | 0.5 [0.3–0.7] | 0.1 [0.0–0.2] | 0.6 [0.4–0.8] | 0.2 [0.1–0.4] | 0.3 [0.2–0.5] |
| Black | 0.1 [0.0–0.3] | 1.3 [0.2–2.4] | 1.1 [0.1–2.1] | 3.1 [1.3–4.9] | 0.1 [0.0–0.3] | 1.1 [0.2–2.0] | 0.7 [0.0–1.6] | 0.6 [0.0–1.7] |
| Asian | - | 1.8 [0.4–3.1] | 0.6 [0.0–1.2] | 3.8 [2.0–5.5] | - | 0.9 [0.0–1.8] | 0.2 [0.0–0.5] | 0.1 [0.0–0.3] |
| Mixed or multiple | 0.4 [0.0–1.2] | 2.1 [0.3–3.8] | 1.7 [0.0–3.5] | 3.8 [1.5–6.1] | - | 0.7 [0.0–1.6] | - | 2.0 [0.0–4.0] |
| Other | - | 1.7 [0.0–4.1] | 1.4 [0.0–4.2] | 3.6 [0.2–6.9] | - | 1.0 [0.0–2.5] | 0.5 [0.0–1.5] | 0.9 [0.0–2.6] |
| Region in England<sup>3</sup> |  |  |  |  |  |  |  |  |
| North | 0.8 [0.3–1.3] | 1.3 [0.8–1.8] | 0.9 [0.4–1.4] | 0.9 [0.5–1.3] | 0.1 [0–0.3] | 0.7 [0.3–1.0] | 0.2 [0.0–0.3] | 0.5 [0.1–0.8] |
| Midlands | 0.2 [0.0–0.4] | 0.9 [0.5–1.4] | 0.5 [0.2–0.8] | 0.5 [0.2–0.7] | 0.1 [0–0.2] | 0.6 [0.3–1.0] | 0.3 [0.1–0.5] | 0.3 [0.0–0.5] |
| South | 0.2 [0.0–0.3] | 1.2 [0.8–1.6] | 0.5 [0.2–0.7] | 1.3 [0.8–1.7] | 0.1 [0–0.3] | 0.6 [0.4–0.9] | 0.3 [0.1–0.5] | 0.5 [0.2–0.7] |
| Vaping status |  |  |  |  |  |  |  |  |
| Current vaping | 0.7 [0.2–1.3] | 3.6 [2.4–4.8] | 1.7 [0.9–2.6] | 3.2 [2.0–4.4] | 0.2 [0–0.5] | 2.0 [1.1–2.9] | 0.7 [0.2–1.2] | 1.3 [0.5–2.1] |
| Not | 0.3 [0.2–0.5] | 0.8 [0.6–1.0] | 0.4 [0.2–0.6] | 0.6 [0.4–0.8] | 0.1 [0–0.2] | 0.4 [0.3–0.6] | 0.2 [0.1–0.3] | 0.3 [0.1–0.4] |

**Table 3. continued**
|  | Prevalence, row % [95%CI] |  |  |  |  |  |  |  |
| --- | --- | --- | --- | --- | --- | --- | --- | --- |
|  | Pipe | Cigar | Cigarillo | Waterpipe | Bidis | Snus | Heated tobacco | Other smokeless tobacco<sup>1</sup> |
| Smoking status |  |  |  |  |  |  |  |  |
| Current smoking<sup>4</sup> | 1.7 [0.9–2.5] | 5.8 [4.4–7.2] | 3.4 [2.3–4.5] | 4.1 [2.9–5.3] | 0.3 [0–0.6] | 1.3 [0.6–1.9] | 1.1 [0.5–1.8] | 1.7 [0.8–2.5] |
| Cigarette smoking | 1.0 [0.4–1.6] | 3.7 [2.5–4.8] | 2.8 [1.7–3.9] | 2.8 [1.7–3.9] | 0.3 [0–0.7] | 1.4 [0.7–2.1] | 1.2 [0.5–1.9] | 1.8 [0.9–2.7] |
| Exclusive non-cigarette tobacco smoking | 9.7 [3.2–16.2] | 30.4 [20.6–40.2] | 10.3 [4.8–15.9] | 19.0 [11.0–27.0] | - | - | 0.6 [0.0–1.8] | - |
| Former smoking | 0.1 [0.0–0.2] | 0.4 [0.1–0.6] | 0.1 [0.0–0.2] | 0.3 [0.1–0.5] | - | 0.8 [0.4–1.3] | 0.2 [0.0–0.4] | 0.4 [0.1–0.7] |
| Never regularly smoked | 0.1 [0.0–0.3] | 0.3 [0.1–0.4] | 0.1 [0.0–0.2] | 0.4 [0.1–0.6] | 0.1 [0–0.3] | 0.4 [0.2–0.5] | 0.0 [0.0–0.1] | 0.1 [0.0–0.1] |
| Nicotine pouch use |  |  |  |  |  |  |  |  |
| No current use | - | - | - | - | - | 0.4 [0.2–0.5] | - | - |
| Current use | - | - | - | - | - | 31.1 [19.3–42.8] | - | - |

<sup>1</sup> Chewing tobacco, paan, gutka, snuff, and dip. <sup>2</sup> Estimates by a more detailed breakdown of ethnicity are provided in **Table S3**. <sup>3</sup> Estimates within each region in the North, Midlands, and South are provided in **Table S5**. <sup>4</sup> Current smoking includes participants who reported smoking cigarettes or other tobacco products. Note: Empty cells indicate where no participants in the subgroup reported use of the product.

**Table 4.**
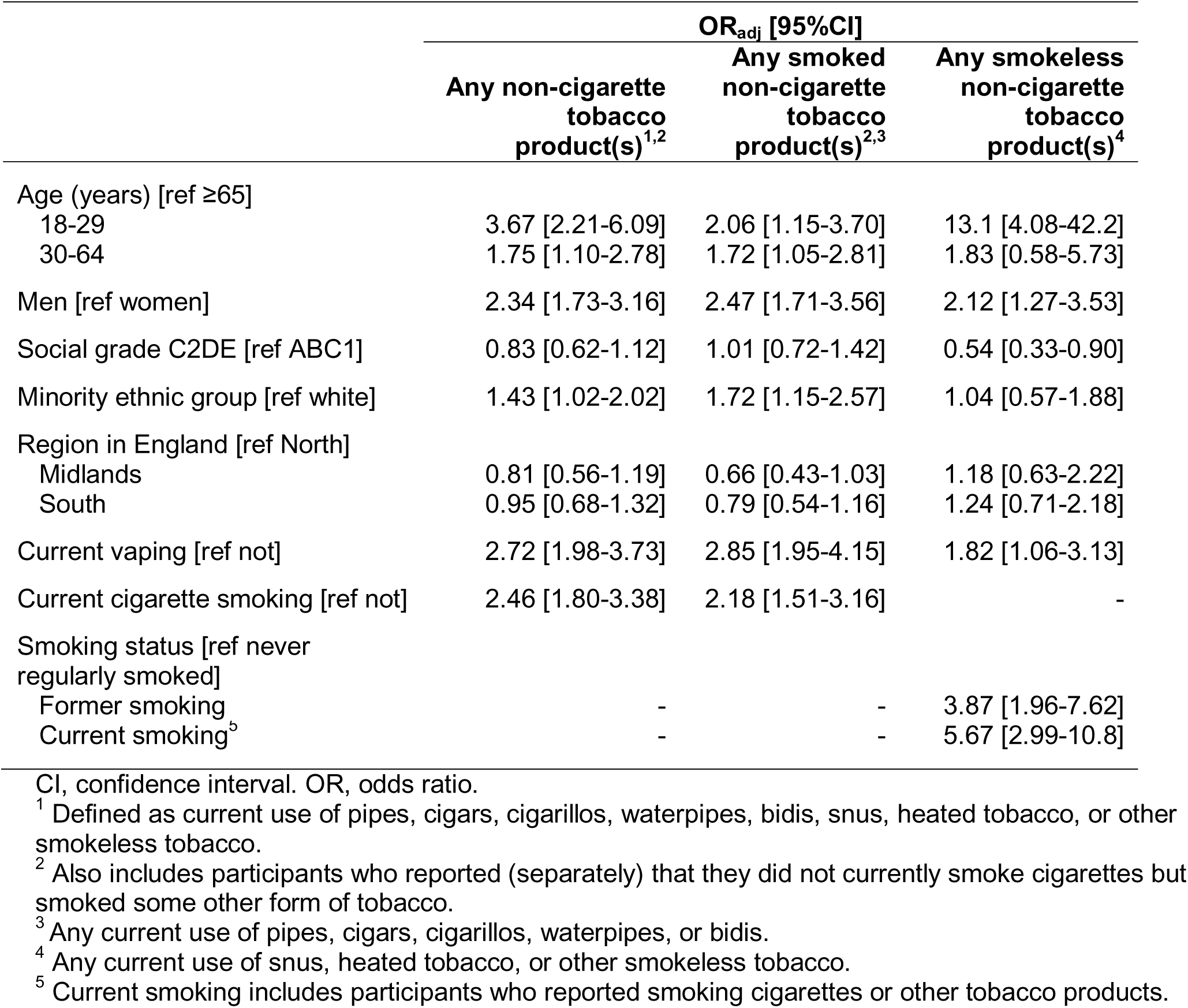
Independent associations of any (i.e., non-exclusive) current use of non-cigarette tobacco with sociodemographic characteristics, vaping status, and smoking status, October 2024 – February 2025 (*n*=8,129; data aggregated across waves)

|  | OR<sub>adj</sub> [95%CI] |  |  |
| --- | --- | --- | --- |
|  | Any non-cigarette tobacco product(s)<sup>1,2</sup> | Any smoked non-cigarette tobacco product(s)<sup>2,3</sup> | Any smokeless non-cigarette tobacco product(s)<sup>4</sup> |
| Age (years) [ref ≥65] |  |  |  |
| 18-29 | 3.67 [2.21-6.09] | 2.06 [1.15-3.70] | 13.1 [4.08-42.2] |
| 30-64 | 1.75 [1.10-2.78] | 1.72 [1.05-2.81] | 1.83 [0.58-5.73] |
| Men [ref women] | 2.34 [1.73-3.16] | 2.47 [1.71-3.56] | 2.12 [1.27-3.53] |
| Social grade C2DE [ref ABC1] | 0.83 [0.62-1.12] | 1.01 [0.72-1.42] | 0.54 [0.33-0.90] |
| Minority ethnic group [ref white] | 1.43 [1.02-2.02] | 1.72 [1.15-2.57] | 1.04 [0.57-1.88] |
| Region in England [ref North] |  |  |  |
| Midlands | 0.81 [0.56-1.19] | 0.66 [0.43-1.03] | 1.18 [0.63-2.22] |
| South | 0.95 [0.68-1.32] | 0.79 [0.54-1.16] | 1.24 [0.71-2.18] |
| Current vaping [ref not] | 2.72 [1.98-3.73] | 2.85 [1.95-4.15] | 1.82 [1.06-3.13] |
| Current cigarette smoking [ref not] | 2.46 [1.80-3.38] | 2.18 [1.51-3.16] | - |
| Smoking status [ref never regularly smoked] |  |  |  |
| Former smoking | - | - | 3.87 [1.96-7.62] |
| Current smoking<sup>5</sup> | - | - | 5.67 [2.99-10.8] |

CI, confidence interval. OR, odds ratio.
<sup>1</sup> Defined as current use of pipes, cigars, cigarillos, waterpipes, bidis, snus, heated tobacco, or other smokeless tobacco.
<sup>2</sup> Also includes participants who reported (separately) that they did not currently smoke cigarettes but smoked some other form of tobacco.
<sup>3</sup> Any current use of pipes, cigars, cigarillos, waterpipes, or bidis.
<sup>4</sup> Any current use of snus, heated tobacco, or other smokeless tobacco.
<sup>5</sup> Current smoking includes participants who reported smoking cigarettes or other tobacco products.

In total, between October 2024 and February 2025, 3.7% reported any current non-cigarette tobacco use, 2.6% reported any current non-cigarette tobacco smoking (i.e., reported exclusive non-cigarette tobacco smoking or any current use of cigars, cigarillos, pipes, waterpipe tobacco, or bidis), and 1.2% reported any current smokeless tobacco use (i.e., any current use of snus, heated tobacco, or other smokeless tobacco) (**Table 2**). The prevalence of dual use of cigarettes and non-cigarette smoked tobacco was 1.2% [0.9–1.4%]; this represented 34.6% [27.3–41.9%] of those who used non-cigarette smoked tobacco. The prevalence of dual use of smoked and smokeless tobacco (including cigarettes) was 0.6% [0.4–0.8%]; this represented 48.7% [37.6–59.8%] of those who used smokeless tobacco.

The use of each individual non-cigarette tobacco product was relatively rare (**Table 3**). Cigars were the most popular smoked tobacco product, used by 1.2% of adults between October 2024 and February 2025 (98 participants), followed by waterpipes (0.9%; 77 participants), cigarillos (0.60%; 48 participants), pipes (0.4%; 28 participants), and bidis (0.1%; 8 participants). Of the smokeless products, 0.6% (53 participants) reported using snus, 0.3% (21 participants) heated tobacco, and 0.4% (29 participants) other smokeless tobacco.

Despite the small numbers of users, we detected some independent differences in product use between population subgroups (**Tables 2-5**). Overall, the odds of using any non-cigarette tobacco product were higher among younger (vs. older) participants, men (vs. women), those from minority (vs. white) ethnic groups, those who reported current vaping (vs. not), and those who reported current cigarette smoking (vs. not). The same differences were observed when we looked specifically at use of smoked non-cigarette tobacco products, but patterns differed slightly for smokeless products: odds were higher among younger (vs. older) participants, men (vs. women), those who reported current vaping (vs. not), and those who reported current or former tobacco smoking (vs. having never regularly smoked), but did not differ significantly by ethnicity.

**Table 5.** Independent associations of any (i.e., non-exclusive) current use of different forms of non-cigarette tobacco with sociodemographic characteristics, vaping status, and smoking status.

|  | OR<sub>adj</sub> [95%CI] |  |  |  |  |  |  |  |
| --- | --- | --- | --- | --- | --- | --- | --- | --- |
|  | Pipe | Cigar | Cigarillo | Waterpipe | Bidis | Snus | Heated tobacco | Other smokeless tobacco<sup>1</sup> |
| Age (years) [ref ≥65] |  |  |  |  |  |  |  |  |
| 18-29 | 0.85 [0.26-2.83] | 1.20 [0.54-2.66] | 1.46 [0.47-4.54] | 12.2 [1.60-92.8] | 3.52 [0.41-29.9] | 61.4 [7.55-499] | 5.10 [0.61-42.8] | 4.52 [0.77-26.5] |
| 30-64 | 0.87 [0.29-2.64] | 1.11 [0.59-2.10] | 1.13 [0.45-2.85] | 6.59 [0.87-49.7] | 0.22 [0.02-2.56] | 3.01 [0.36-25.3] | 2.81 [0.39-20.3] | 0.82 [0.15-4.61] |
| Men [ref women] | 3.19 [1.07-9.52] | 12.5 [4.84-32.4] | 2.93 [1.25-6.86] | 1.67 [0.97-2.88] | 0.71 [0.14-3.61] | 3.27 [1.64-6.51] | 0.88 [0.33-2.36] | 2.62 [0.96-7.12] |
| Social grade C2DE [ref ABC1] | 1.34 [0.52-3.45] | 0.67 [0.42-1.08] | 0.88 [0.45-1.73] | 0.66 [0.36-1.22] | 0.48 [0.05-4.72] | 0.37 [0.17-0.81] | 0.38 [0.12-1.19] | 1.20 [0.52-2.79] |
| Minority ethnic group [ref white] | 0.16 [0.02-1.23] | 1.51 [0.84-2.71] | 1.97 [0.88-4.39] | 6.66 [3.49-12.7] | 0.24 [0.02-3.20] | 0.79 [0.37-1.69] | 1.22 [0.32-4.68] | 1.39 [0.48-4.04] |
| Region in England [ref North] |  |  |  |  |  |  |  |  |
| Midlands | 0.22 [0.07-0.70] | 0.82 [0.44-1.52] | 0.67 [0.27-1.63] | 0.48 [0.21-1.09] | 0.65 [0.11-3.92] | 1.08 [0.48-2.46] | 2.09 [0.62-7.08] | 0.46 [0.12-1.68] |
| South | 0.21 [0.07-0.61] | 0.91 [0.53-1.55] | 0.49 [0.22-1.11] | 1.10 [0.56-2.14] | 0.99 [0.16-6.31] | 0.86 [0.43-1.75] | 1.82 [0.53-6.17] | 0.94 [0.35-2.56] |
| Current vaping [ref not] | 1.54 [0.53-4.47] | 3.39 [2.03-5.64] | 2.15 [0.99-4.65] | 4.84 [2.39-9.80] | 1.74 [0.42-7.22] | 1.87 [0.85-4.11] | 1.39 [0.50-3.88] | 1.91 [0.78-4.66] |
| Current cigarette smoking [ref not] | 1.83 [0.67-5.02] | 3.41 [2.07-5.63] | 11.0 [5.58-21.8] | 2.81 [1.43-5.53] | 2.58 [0.62-10.7] | - | - | - |
| Smoking status [ref never regularly smoked] |  |  |  |  |  |  |  |  |
| Former smoking | - | - | - | - | - | 3.25 [1.32-8.04] | 5.36 [0.88-32.9] | 7.26 [1.68-31.4] |
| Current smoking<sup>2</sup> | - | - | - | - | - | 2.09 [0.92-4.71] | 27.9 [5.56-139] | 16.3 [3.83-69.3] |

CI, confidence interval. OR<sub>adj</sub>, adjusted odds ratio.
Each column presents data from a separate model with use of the stated product as the dependent variable and age, gender, occupational social grade, ethnicity, region, and cigarette smoking (for analyses of smoked tobacco product use) or smoking status (for analyses of smokeless tobacco product use) as independent variables.
<sup>1</sup> Chewing tobacco (e.g. paan, gutka), snuff, and dip. <sup>2</sup> Current smoking includes participants who reported smoking cigarettes or other tobacco products.

However, these patterns varied between specific products (**Table 3**, **Table 5**). The odds of using cigars, cigarillos, and pipes were higher among men (vs. women) and among those who reported current cigarette smoking (vs. not). The odds of using cigars were also higher among those who reported current vaping (vs. not) and the odds of using pipes were also higher among those in the North of England. The odds of using waterpipes were higher among younger (vs. older) participants, those from minority (vs. white) ethnic groups, and those who reported current vaping (vs. not). The odds of using snus were higher among younger (vs. older) participants, those from more (vs. less) advantaged social grades, and those who reported former tobacco smoking (vs. having never regularly smoked tobacco). Snus use was also much more common among those who reported current use of tobacco-free nicotine pouches than those who did not. The odds of using heated tobacco were higher among those who reported current (vs. never regular) tobacco smoking, while the odds of other smokeless tobacco use were higher among those who reported current or former (vs. never regular) tobacco smoking.

## Discussion

The findings of this study provide up-to-date insights into patterns of non-cigarette tobacco use among adults in England and how they vary across different population subgroups.

The prevalence of exclusive non-cigarette tobacco smoking (i.e., the proportion of adults who do not currently smoke cigarettes but use some other form of smoked tobacco) fluctuated over the study period, initially increasing between 2020 and 2022 before declining by 2025. This suggests that the rising trend in the exclusive use of non-cigarette smoked tobacco, particularly during the early 2020s, that we previously reported^3^ has not been sustained. The reason for this uptick is not clear, but plausibly it may reflect increased experimentation during the COVID-19 pandemic period and associated lockdowns or post-pandemic socialising, while the later decline could indicate a return to pre-pandemic patterns or substitution with other nicotine products, such as vaping (which became much more popular in England from mid-2021 onwards^5^). Notably, these fluctuations were most pronounced among younger adults and those who vaped, consistent with evidence that these groups are more likely to experiment with alternative tobacco forms.^26,27^ Our previous analysis suggested prevalence of exclusive non-cigarette tobacco smoking rose sharply at the start of the pandemic,^3^ so although current levels (2025: 1.2%) are now similar to the start of the pandemic, they remain higher than they were pre-pandemic (2013; 0.4%). In addition, these estimates only capture exclusive use: in recent survey waves, the prevalence of any vs. exclusive non-cigarette tobacco smoking was around twice as high (2.6% vs. 1.2%). Including the use of smokeless tobacco products, we estimated the prevalence of any non-cigarette tobacco product use in late 2024/early 2025 to be 3.7%. Extrapolated to the general population, this equates to approximately 1.7 million people (45.7 million adults ≥18y in England^28^ x 3.7%) using any smoked or smokeless non-cigarette tobacco.

Although the overall use of specific non-cigarette tobacco products was relatively rare, we found evidence of sociodemographic clustering consistent with previous research.^12,13,29^ Cigars were the most commonly used smoked product (particularly among men), followed closely by waterpipes. Use of cigars, cigarillos, and pipes was more prevalent among men and those who currently smoke, reflecting traditional gendered patterns of tobacco use.^29^ Use of waterpipes was more prevalent among younger adults, some ethnic minority groups, and those who vaped. These findings echo previous reports that waterpipe use is particularly appealing to young people and some minority ethnic communities^12,13^ due to its perceived lower harm and its integration into social and cultural practices – for instance, waterpipe use often occurs in group settings in bars, restaurants, and cafes.^11,17,30^ Bidis were rarely used, likely reflecting limited availability and use within specific (i.e., South Asian) communities.^31^

Among smokeless products, snus was the most commonly used, followed by other forms of smokeless tobacco and heated tobacco products. Snus use was more prevalent among younger adults, those from more advantaged social grades, and people who formerly smoked tobacco. This may reflect experimentation with other nicotine products among those seeking (less harmful) alternatives to smoking,^32^ or crossover from users of tobacco-free nicotine pouches – as suggested by our exploratory analysis (nicotine pouches are also more popular among younger adults^4^).

However, there may also have been some misreporting of tobacco-free nicotine pouch use as snus use, given the products are similar in appearance and method of use and some major media outlets have referred to nicotine pouches as snus.^33^ As has been observed in other studies,^34^ use of heated tobacco was more strongly associated with current smoking, suggesting it may be used more commonly as a complement or transition product rather than a replacement. However, this pattern might also reflect the fact that heated tobacco products are a relatively new product category, and it is possible their use may increase among those who formerly smoked over time as more people transition from smoking to heated tobacco use, although there is not strong evidence for this as yet.^34^ Use of other smokeless tobacco products (e.g., paan, gutka, snuff) was more prevalent among those who reported current or former smoking. However, while previous literature suggests these products are more commonly used in South Asian communities,^14,15^ we did not detect differences in the use of other smokeless tobacco products by ethnicity, which may reflect this category encompassing a range of different products, the low prevalence of use in the analysed sample, and the small numbers of participants from specific ethnic minority groups.

Key strengths of this study include the large, nationally representative sample and the granularity of product-specific data. The inclusion of both smoked and smokeless tobacco products allows for a comprehensive picture of tobacco use in England. However, there are also limitations. The reliance on self-reported data without verification by asking participants to display the product(s) they use may introduce some degree of reporting bias (e.g., if participants reported use of nicotine pouches as snus). Interviews were only conducted in English, so the sample may have excluded people who did not speak English as their first language or struggled to understand English over the telephone. This may particularly affect those from South Asian communities,^35^ which might explain why we did not detect anticipated differences in product use by ethnicity and may have resulted in us underestimating the prevalence of use of certain products. There were only a small number of users of each non-cigarette tobacco product between October 2024 and February 2025, resulting in imprecision in prevalence estimates (indicated by wide confidence intervals) and limited statistical power to detect subgroup differences. In addition, small numbers in minority ethnic groups meant we lacked statistical power to examine differences between these groups. Further research with oversampling of minority ethnic groups would be useful to gain more insight. Finally, the grouping of diverse smokeless tobacco products into a single category limited insights into the specific products being used by different population subgroups.

Nonetheless, this study underscores the importance of monitoring non-cigarette tobacco use as part of broader tobacco control efforts. Collectively, non-cigarette tobacco products are used by approximately 1.7 million adults in England, many of whom do not smoke cigarettes so will not be captured by surveys that focus only on cigarette smoking (e.g., the Annual Population Survey^36^) or by tobacco cessation services and outreach programmes. Further, rates are higher among specific subgroups – such as younger adults, men, ethnic minority groups, and those who currently smoke. Public health strategies should consider these disparities and ensure that messaging about the risks of non-cigarette tobacco use reaches all populations effectively to reduce tobacco-related health inequalities, and that stop smoking services do not focus only on cigarettes but provide support for cessation of the wide range of tobacco products. Given the evolving tobacco landscape, continued surveillance and research into the motivations for using non-cigarette tobacco products will be important for informing future policy and harm reduction strategies. Monitoring should capture both exclusive and co-use patterns, as risks may differ, and consider using visual aids or examples to clarify product categories to improve data quality and public understanding.

## Data Availability

All data produced in the present study are available upon reasonable request to the authors

## Declarations

### Data availability

The data use in these analyses are available on Open Science Framework (https://osf.io/7vhk5/), with age provided in bands to preserve anonymity.

### Ethics approval

Ethical approval for the STS was granted originally by the UCL Ethics Committee (ID 0498/001). Participants provide informed consent to take part in the study, and all methods are carried out in accordance with relevant regulations. The data are not collected by UCL and are anonymised when received by UCL.

## References

1. World Health Organization. Tobacco. 2023. https://www.who.int/news-room/fact-sheets/detail/tobacco#:∼:text=Over%2080%25%20of%20the%20world’s,(WHO%20FCTC)%20in%202003. (accessed 23 Apr2025).

2 Foulds J, Ramstrom L, Burke M, Fagerström K. Effect of smokeless tobacco (snus) on smoking and public health in Sweden. Tob Control 2003; 12: 349–359.

3 Jackson SE, Shahab L, Brown J. Trends in Exclusive Non-Cigarette Tobacco Smoking in England: A Population Survey 2013–2023. Nicotine Tob Res 2024; : ntae021.

4 Tattan-Birch H, Jackson SE, Dockrell M, Brown J. Tobacco-free Nicotine Pouch Use in Great Britain: A Representative Population Survey 2020–2021. Nicotine Tob Res 2022; : ntac099.

5 Jackson SE, Tattan-Birch H, Shahab L, Brown J. Trends in long term vaping among adults in England, 2013-23: population based study. BMJ 2024; **386**: e079016.

6 Hajat C, Stein E, Ramstrom L, Shantikumar S, Polosa R. The health impact of smokeless tobacco products: a systematic review. Harm Reduct J 2021; 18: 123.

7 Tattan-Birch H, Hartmann-Boyce J, Kock L, Simonavicius E, Brose L, Jackson S et al. Heated tobacco products for smoking cessation and reducing smoking prevalence. Cochrane Database Syst Rev 2022. doi:10.1002/14651858.CD013790.pub2.

8 Baker F, Ainsworth SR, Dye JT, Crammer C, Thun MJ, Hoffmann D et al. Health Risks Associated With Cigar Smoking. JAMA 2000; 284: 735–740.

9 Waziry R, Jawad M, Ballout RA, Al Akel M, Akl EA. The effects of waterpipe tobacco smoking on health outcomes: an updated systematic review and meta-analysis. Int J Epidemiol 2017; 46: 32–43.

10 Valen H, Becher R, Vist GE, Holme JA, Mdala I, Elvsaas I-KØ et al. A systematic review of cancer risk among users of smokeless tobacco (Swedish snus) exclusively, compared with no use of tobacco. Int J Cancer 2023; **153**: 1942–1953.

11 Akl EA, Jawad M, Lam WY, Co CN, Obeid R, Irani J. Motives, beliefs and attitudes towards waterpipe tobacco smoking: a systematic review. Harm Reduct J 2013; 10: 12.

12 Grant A, Morrison R, Dockrell MJ. Prevalence of Waterpipe (Shisha, Narghille, Hookah) Use Among Adults in Great Britain and Factors Associated With Waterpipe Use: Data From Cross-sectional Online Surveys in 2012 and 2013. Nicotine Tob Res 2014; **16**: 931–938.

13 Jackson D, Aveyard P. Waterpipe smoking in students: Prevalence, risk factors, symptoms of addiction, and smoke intake. Evidence from one British university. BMC Public Health 2008; 8: 174.

14 West R, McNeill A, Raw M. Smokeless tobacco cessation guidelines for health professionals in England. Br Dent J 2004; 196: 611–618.

15 Longman JM, Pritchard C, McNeill A, Csikar J, Croucher RE. Accessibility of chewing tobacco products in England. J Public Health 2010; 32: 372–378.

16 Moles DR, Fedele S, Speight PM, Porter SR, Silva I dos S. Oral and pharyngeal cancer in South Asians and non-South Asians in relation to socioeconomic deprivation in South East England. Br J Cancer 2008; 98: 633–635.

17. Rennie D. Cancer Research UK’s February 2023 Cancer Awareness measure (CAM). 2023.

18 Li L, Borland R, Cummings KM, Gravely S, Quah ACK, Fong GT et al. Patterns of Non-Cigarette Tobacco and Nicotine Use Among Current Cigarette Smokers and Recent Quitters: Findings From the 2020 ITC Four Country Smoking and Vaping Survey. Nicotine Tob Res 2021; 23: 1611–1616.

19 Hammond D, Reid JL, Burkhalter R, D’Mello K. Trends in smoking and vaping among young people: findings from the ITC Youth & Young Adult Tobacco and Vaping Survey, 2017-2023. 2024 https://davidhammond.ca/wp-content/uploads/2025/02/ITC_Youth_Report_2017-2023.pdf (accessed 23 Apr2025).

20. HM Revenue & Customs. Tobacco statistics commentary January 2025. GOV.UK. 2025. https://www.gov.uk/government/statistics/tobacco-bulletin/tobacco-statistics-commentary-april-2023--2 (accessed 22 May2024).

21 Fidler JA, Shahab L, West O, Jarvis MJ, McEwen A, Stapleton JA et al. ‘The smoking toolkit study’: a national study of smoking and smoking cessation in England. BMC Public Health 2011; 11: 479.

22 Kock L, Shahab L, Moore G, Beard E, Bauld L, Reid G, et al. Protocol for expansion of an existing national monthly survey of smoking behaviour and alcohol use in England to Scotland and Wales: The Smoking and Alcohol Toolkit Study. Wellcome Open Res 2021; 6: 67.

23 Jackson SE, Beard E, Kujawski B, Sunyer E, Michie S, Shahab L et al. Comparison of Trends in Self-reported Cigarette Consumption and Sales in England, 2011 to 2018. JAMA Netw Open 2019; **2**: e1910161.

24. National Readership Survey. Social Grade. http://www.nrs.co.uk/nrs-print/lifestyle-and-classification-data/social-grade/ (accessed 3 Feb2019).

25. Office for National Statistics. Ethnic group classifications: Census 2021. 2023.https://www.ons.gov.uk/census/census2021dictionary/variablesbytopic/ethnicgroupnationalidentitylanguageandreligionvariablescensus2021/ethnicgroup/classifications (accessed 23 Apr2025).

26 Auf R, Trepka MJ, Selim M, Taleb ZB, Rosa MDL, Bastida E et al. E-cigarette use is associated with other tobacco use among US adolescents. Int J Public Health 2019; 64: 125–134.

27 Leventhal AM, Strong DR, Kirkpatrick MG, Unger JB, Sussman S, Riggs NR et al. Association of Electronic Cigarette Use With Initiation of Combustible Tobacco Product Smoking in Early Adolescence. JAMA 2015; 314: 700–707.

28. Office for National Statistics. Population estimates for the UK, England, Wales, Scotland, and Northern Ireland: mid-2022. 2024. https://www.ons.gov.uk/peoplepopulationandcommunity/populationandmigration/populationestimates/bulletins/annualmidyearpopulationestimates/mid2022 (accessed 5 Jun2024).

29 Peters SAE, Huxley RR, Woodward M. Do smoking habits differ between women and men in contemporary Western populations? Evidence from half a million people in the UK Biobank study. BMJ Open 2014; 4: e005663.

30 Arshad A, Matharoo J, Arshad E, Sadhra SS, Norton-Wangford R, Jawad M. Knowledge, attitudes, and perceptions towards waterpipe tobacco smoking amongst college or university students: a systematic review. BMC Public Health 2019; 19: 439.

31 Pednekar MS, Gupta PC, Yeole BB, Hébert JR. Association of tobacco habits, including bidi smoking, with overall and site-specific cancer incidence: results from the Mumbai cohort study. Cancer Causes Control 2011; 22: 859–868.

32 Gartner CE, Hall WD, Vos T, Bertram MY, Wallace AL, Lim SS. Assessment of Swedish snus for tobacco harm reduction: an epidemiological modelling study. The Lancet 2007; 369: 2010– 2014.

33. Roberts G. I gave up vaping. Now I’m addicted to snus. The Times. 2025. https://www.thetimes.com/life-style/health-fitness/article/vaping-alternative-snus-5vvsv02qh (accessed 9 May2025).

34 Scala M, Dallera G, Gorini G, Achille J, Havermans A, Neto C et al. Patterns of use of heated tobacco products: a comprehensive systematic review. J Epidemiol 2025. doi:10.2188/jea.JE20240189.

35. English language skills. GOV.UK. 2018.https://www.ethnicity-facts-figures.service.gov.uk/uk-population-by-ethnicity/demographics/english-language-skills/latest/ (accessed 23 Apr2025).

36. Office for National Statistics. Adult smoking habits in the UK: 2023. 2024 https://www.ons.gov.uk/peoplepopulationandcommunity/healthandsocialcare/healthandlifeexpectancies/bulletins/adultsmokinghabitsingreatbritain/2023 (accessed 1 Oct2024).

