## Supplementary material for "Prevalence and patterns of different types of non-cigarette tobacco use in England: a population study": Table S1

**Table S1.** Sample characteristics

| | Apr 2020 – Feb 2025 | | Oct 2024 – Feb 2025 | |
|---|---|---|---|---|
| | N[1] | Weighted %[2] | N[1] | Weighted %[2] |
| All adults | 94,918 | 100.0 | 8,129 | 100.0 |
| Age group (years) | | | | |
| 18-29 | 16,453 | 18.9 | 1,463 | 19.6 |
| 30-64 | 54,136 | 58.0 | 4,611 | 57.7 |
| ≥65 | 24,329 | 23.1 | 2,055 | 22.8 |
| *Missing* | - | - | - | - |
| Gender | | | | |
| Men | 46,547 | 48.7 | 4,100 | 48.8 |
| Women | 47,389 | 50.6 | 3,913 | 50.4 |
| In another way | 694 | 0.7 | 72 | 0.8 |
| *Missing* | 288 | - | 44 | - |
| Occupational social grade | | | | |
| ABC1 (more advantaged) | 63,359 | 56.2 | 5,738 | 56.4 |
| C2DE (less advantaged) | 31,559 | 43.8 | 2,391 | 43.6 |
| *Missing* | - | - | - | - |
| Ethnicity[3] | | | | |
| White | 72,819 | 85.5 | 6,660 | 84.6 |
| Black | 4,502 | 4.9 | 493 | 5.5 |
| Asian | 4,835 | 5.5 | 495 | 5.5 |
| Mixed/multiple | 2,480 | 2.7 | 274 | 3.0 |
| Other | 1,271 | 1.4 | 130 | 1.4 |
| *Missing* | 9,011 | - | 77 | - |
| Region in England | | | | |
| North | 25,991 | 27.7 | 2,190 | 27.6 |
| Midlands | 28,451 | 30.3 | 2,380 | 30.1 |
| South | 40,476 | 42.1 | 3,559 | 42.3 |
| *Missing* | - | - | - | - |
| Vaping status | | | | |
| Current vaping | 8,612 | 9.9 | 965 | 13.4 |
| Not | 86,306 | 90.1 | 7,164 | 86.6 |
| *Missing* | - | - | - | - |
| Smoking status | | | | |
| Current smoking[4] | 14,466 | 16.5 | 1,138 | 15.4 |
| Cigarette smoking | 12,906 | 14.8 | 1,037 | 14.1 |
| Exclusive non-cigarette smoking | 1,560 | 1.7 | 101 | 1.3 |
| Former smoking | 25,636 | 26.4 | 2,370 | 28.8 |
| Never regularly smoked | 54,816 | 57.1 | 4,621 | 55.8 |
| *Missing* | - | - | - | - |

CI, confidence interval.

[1] Unweighted sample size.

[2] Valid percentages shown.

[3] Ethnicity was not assessed in April – August 2020; the number of missing cases includes participants surveyed in these waves.

[4] Current smoking includes participants who reported smoking cigarettes or other tobacco products.

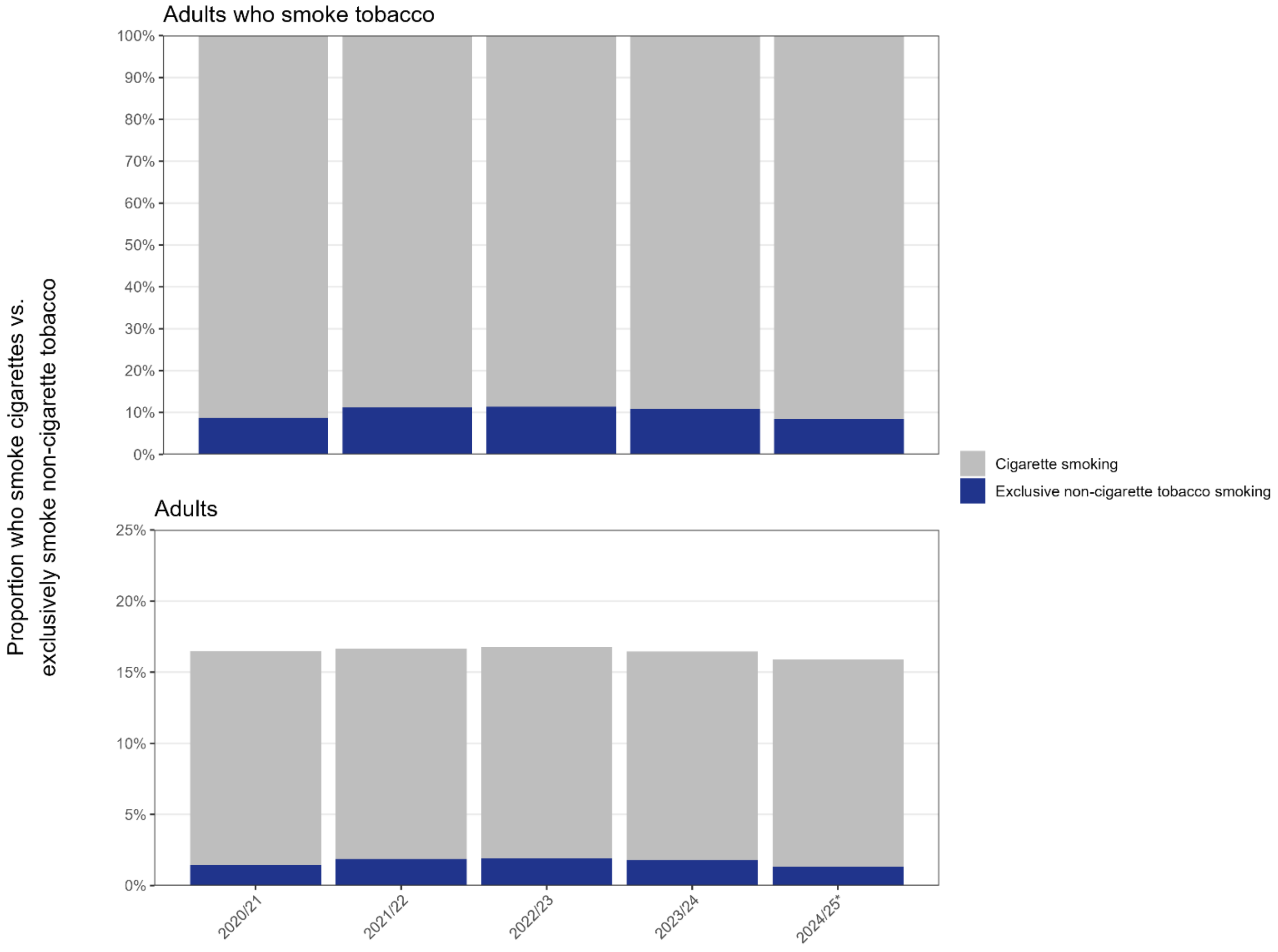


**Figure S1. Prevalence of cigarette smoking and exclusive non-cigarette smoking among adults (≥18y) in England, 2020 to 2025**

Panels show weighted data aggregated by year, among adults who smoke tobacco and among all adults. Bars represent the proportions smoking cigarettes vs. exclusively smoking non-cigarette tobacco. Data are aggregated across 12-month periods (April to March). *Data for 2024/25 are based on April to February only.

**Table S2.** Ethnic differences in any current use of non-cigarette tobacco among adults in England, October 2024 – February 2025 (*n*=8,129; data aggregated across waves)

| | | Prevalence, row % [95%CI] | | |
|---|---|---|---|---|
| | ***N*[1]** | **Any non-cigarette tobacco product(s)[2,3]** | **Any smoked non-cigarette tobacco product(s)[3,4]** | **Any smokeless non-cigarette tobacco product(s)[5]** |
| Ethnicity | | | | |
| White British | 6,114 | 3.1 [2.7–3.6] | 2.3 [1.8–2.7] | 1.0 [0.7–1.3] |
| White Irish | 102 | 2.9 [0.0–7.1] | 2.9 [-1.4–7.1] | - |
| White Gypsy/Traveller/Other | 444 | 4.8 [2.5–7.2] | 2.4 [0.5–4.3] | 2.6 [1.2–4.0] |
| Mixed White/Black Caribbean | 71 | 6.6 [0.0–13.1] | 5.1 [0.0–11] | 1.5 [0.0–4.4] |
| Mixed White/Black African | 41 | 5.3 [0.0–15.7] | - | 5.3 [0.0–15.7] |
| Mixed White/Asian | 68 | 4.7 [0.0–9.5] | 4.7 [0.0–9.5] | - |
| Mixed Other | 94 | 4.8 [0.5–9.1] | 3.0 [0.0–6.4] | 3.1 [0.0–6.7] |
| Asian Indian | 224 | 7.1 [3.2–11.0] | 6.4 [2.6–10.1] | 1.6 [0.0–3.5] |
| Asian Pakistani | 133 | 6.5 [2.4–10.7] | 5.6 [1.6–9.6] | 0.9 [0.0–2.2] |
| Asian Bangladeshi | 41 | - | - | - |
| Asian Chinese/Other | 97 | 1.0 [0.0–3.1] | - | 1.0 [0.0–3.1] |
| Black African | 300 | 5.3 [2.3–8.3] | 2.0 [0.3–3.8] | 3.3 [0.7–5.8] |
| Black Caribbean | 149 | 7.3 [2.1–12.5] | 6.8 [1.7–11.9] | 0.9 [0.4–2.1] |
| Black Other | 44 | 10.6 [1.0–20.3] | 10.6 [1.0–20.3] | - |
| Arab | 35 | 9.8 [0.0–19.9] | 4.3 [0.0–10.5] | 5.5 [0.0–13.7] |
| Other | 95 | 5.4 [0.2–10.7] | 4.1 [0.0–9.1] | 1.3 [0.0–3.2] |

[1] Unweighted sample size.
[2] Defined as current use of pipes, cigars, cigarillos, waterpipes, bidis, snus, heated tobacco, or other smokeless tobacco.
[3] Also includes participants who reported (separately) that they did not currently smoke cigarettes but smoked some other form of tobacco.
[4] Any current use of pipes, cigars, cigarillos, waterpipes, or bidis.
[5] Any current use of snus, heated tobacco, or other smokeless tobacco.
Note: Empty cells indicate where no participants in the subgroup reported use of the product.

**Table S3.** Ethnic differences in any current use of different forms of non-cigarette tobacco among adults in England, October 2024 – February 2025 (*n*=8,129; data aggregated across waves)

| | Prevalence, % [95%CI] | | | | | | | |
|---|---|---|---|---|---|---|---|---|
| | **Pipe** | **Cigar** | **Cigarillo** | **Waterpipe** | **Bidis** | **Snus** | **Heated tobacco** | **Other smokeless tobacco[1]** |
| Ethnicity | | | | | | | | |
| White British | 0.3 [0.2–0.5] | 1.1 [0.8–1.3] | 0.5 [0.3–0.7] | 0.4 [0.2–0.6] | 0.1 [0.0–0.2] | 0.6 [0.4–0.8] | 0.2 [0.0–0.3] | 0.3 [0.2–0.5] |
| White Irish | - | 2.9 [0.0–7.1] | 0.8 [0.0–2.5] | - | - | - | - | - |
| White Gypsy/Traveller/Other | 1.4 [0.0–2.9] | 0.4 [0.0–1.0] | 0.2 [0.0–0.6] | 1.7 [0.0–3.3] | 0.2 [0.0–0.6] | 0.8 [0.0–1.7] | 1.4 [0.4–2.5] | 0.6 [0.0–1.3] |
| Mixed White/Black Caribbean | - | 2.1 [0.0–6.2] | 3.2 [0.0–7.9] | 1.3 [0.0–4.0] | - | - | - | 1.5 [0.0–4.4] |
| Mixed White/Black African | - | - | - | 4.1 [0.0–9.9] | - | - | - | 5.3 [0.0–15.7] |
| Mixed White/Asian | - | 3.8 [0.0–8.4] | 1.8 [0.0–5.3] | 3.9 [0.0–8.4] | - | - | - | - |
| Mixed Other | 1.3 [0.0–3.7] | 1.7 [0.0–4.1] | 1.3 [0.0–3.7] | 5.7 [0.4–11.1] | - | 2.0 [0.0–5.0] | - | 2.3 [0.0–5.6] |
| Asian Indian | - | 3.6 [0.6–6.6] | 0.8 [0.0–2.0] | 3.2 [0.7–5.6] | - | 1.3 [0.0–3.3] | - | 0.2 [0.0–0.7] |
| Asian Pakistani | - | 0.7 [0.0–2.1] | 0.7 [0.0–2.2] | 6.8 [2.6–11.0] | - | 0.4 [0.0–1.1] | 0.5 [0.0–1.6] | - |
| Asian Bangladeshi | - | - | - | 1.1 [0.0–3.3] | - | - | - | - |
| Asian Chinese/Other | - | - | - | 1.9 [0.0–5.7] | - | 1.0 [0.0–3.1] | - | - |
| Black African | - | 0.6 [0.0–1.4] | 1.2 [0.0–2.7] | 3.2 [0.8–5.7] | - | 1.4 [0.1–2.7] | 1.1 [0.0–2.5] | 0.8 [0.0–2.5] |
| Black Caribbean | 0.4 [0.0–1.2] | 2.5 [0.0–5.5] | 1.3 [0.0–2.7] | 3.0 [0.0–6.1] | 0.41 [0.0–1.2] | 0.9 [0.0–2.1] | - | 0.4 [0.0–1.2] |
| Black Other | - | 2.3 [0.0–7.0] | - | 2.3 [0.0–7.0] | - | - | - | - |
| Arab | - | - | - | 9.7 [0.0–19.7] | - | 2.1 [0.0–6.5] | - | 3.5 [0.0–10.7] |
| Other | - | 2.2 [0.0–5.5] | 1.9 [0.0–5.6] | 1.5 [0.0–4.6] | - | 0.7 [0.0–2.0] | 0.7 [0.0–2.0] | - |

[1] Chewing tobacco, paan, gutka, snuff, and dip.
Note: Empty cells indicate where no participants in the subgroup reported use of the product.

**Table S4.** Regional differences in any current use of non-cigarette tobacco among adults in England, October 2024 – February 2025 (*n*=8,129; data aggregated across waves)

| | | Prevalence, row % [95%CI] | | |
|---|---|---|---|---|
| | *N*[1] | Any non-cigarette tobacco product(s)[2,3] | Any smoked non-cigarette tobacco product(s)[3,4] | Any smokeless non-cigarette tobacco product(s)[5] |
| Region in England | | | | |
| North East | 363 | 2.7 [1.0–4.4] | 2.3 [0.7–3.8] | 0.6 [0.0–1.2] |
| North West | 1,050 | 3.8 [2.4–5.1] | 2.9 [1.7–4.2] | 1.3 [0.6–2.1] |
| Yorkshire and the Humber | 777 | 4.7 [3.0–6.4] | 3.8 [2.2–5.4] | 1.0 [0.3–1.6] |
| East Midlands | 691 | 3.7 [2.0–5.3] | 2.9 [1.4–4.5] | 1.0 [0.1–1.8] |
| West Midlands | 771 | 2.4 [1.3–3.5] | 1.7 [0.8–2.6] | 0.8 [0.1–1.5] |
| East of England | 918 | 3.3 [2.0–4.6] | 1.6 [0.7–2.4] | 1.8 [0.7–2.8] |
| London | 1,378 | 4.5 [3.2–5.7] | 3.2 [2.1–4.2] | 1.5 [0.8–2.2] |
| South East | 1,379 | 3.3 [2.3–4.4] | 2.6 [1.6–3.5] | 0.9 [0.4–1.4] |
| South West | 802 | 3.7 [2.3–5.1] | 1.9 [1.0–2.8] | 1.8 [0.7–2.9] |

[1] Unweighted sample size.
[2] Defined as current use of pipes, cigars, cigarillos, waterpipes, bidis, snus, heated tobacco, or other smokeless tobacco.
[3] Also includes participants who reported (separately) that they did not currently smoke cigarettes but smoked some other form of tobacco.
[4] Any current use of pipes, cigars, cigarillos, waterpipes, or bidis.
[5] Any current use of snus, heated tobacco, or other smokeless tobacco.
Note: Empty cells indicate where no participants in the subgroup reported use of the product.

**Table S5.** Regional differences in any current use of different forms of non-cigarette tobacco among adults in England, October 2024 – February 2025 (*n*=8,129; data aggregated across waves)

| | Prevalence, % [95%CI] | | | | | | | |
|---|---|---|---|---|---|---|---|---|
| | Pipe | Cigar | Cigarillo | Waterpipe | Bidis | Snus | Heated tobacco | Other smokeless tobacco[1] |
| Region in England | | | | | | | | |
| North East | 0.3 [0.0–0.8] | 1.6 [0.2–3.0] | 0.3 [0.0–0.8] | 0.3 [0.0–0.8] | 0.1 [0.0–0.4] | 0.6 [0.0–1.2] | - | 0.1 [0.0–0.4] |
| North West | 0.9 [0.1–1.7] | 1.2 [0.5–1.9] | 0.8 [0.1–1.5] | 1.1 [0.3–1.8] | 0.2 [0.0–0.5] | 0.7 [0.2–1.2] | 0.2 [0.0–0.4] | 0.8 [0.2–1.5] |
| Yorkshire and the Humber | 0.9 [0.1–1.7] | 1.4 [0.5–2.3] | 1.2 [0.3–2.2] | 0.9 [0.3–1.6] | - | 0.7 [0.1–1.3] | 0.2 [0.0–0.5] | 0.1 [0.0–0.4] |
| East Midlands | - | 1.5 [0.4–2.5] | 0.8 [0.1–1.4] | 0.2 [0.0–0.5] | - | 0.5 [0.0–1.0] | 0.5 [0.0–1.1] | - |
| West Midlands | 0.3 [0.0–0.7] | 1.0 [0.3–1.7] | 0.4 [0.0–0.9] | 0.8 [0.1–1.4] | 0.2 [0.0–0.5] | 0.4 [0.0–0.8] | - | 0.5 [0.0–1.0] |
| East of England | 0.2 [0.0–0.5] | 0.5 [0.0–0.9] | 0.3 [0.0–0.7] | 0.4 [0.0–0.8] | - | 1.0 [0.2–1.9] | 0.4 [0.0–0.9] | 0.3 [0.0–0.7] |
| London | 0.1 [0.0–0.3] | 1.3 [0.7–2.0] | 0.6 [0.1–1.0] | 2.3 [1.3–3.3] | - | 0.6 [0.2–1.0] | 0.3 [0.0–0.5] | 0.6 [0.1–1.1] |
| South East | 0.3 [0.0–0.6] | 1.3 [0.7–1.8] | 0.3 [0.0–0.6] | 0.8 [0.3–1.3] | 0.4 [0.0–0.8] | 0.4 [0.1–0.8] | 0.3 [0.0–0.5] | 0.3 [0.0–0.6] |
| South West | 0.1 [0.0–0.3] | 0.9 [0.3–1.6] | 0.6 [0.1–1.0] | 0.3 [0.0–0.7] | - | 0.9 [0.3–1.5] | 0.4 [0.0–1.1] | 0.5 [0.0–1.1] |

[1] Chewing tobacco, paan, gutka, snuff, and dip.

Note: Empty cells indicate where no participants in the subgroup reported use of the product.

**Table S6.** Unadjusted associations of any current use of non-cigarette tobacco with sociodemographic characteristics, vaping status, and smoking status

| | OR [95%CI] | | |
|---|---|---|---|
| | Any non-cigarette tobacco product(s)[1,2] | Any smoked non-cigarette tobacco product(s)[2,3] | Any smokeless non-cigarette tobacco product(s)[4] |
| Age (years) [ref ≥65] | | | |
| 18-29 | 5.98 [3.79-9.42] | 3.63 [2.18-6.06] | 15.5 [5.21-46.1] |
| 30-64 | 2.44 [1.57-3.78] | 2.40 [1.51-3.80] | 2.49 [0.82-7.52] |
| Men [ref women] | 2.48 [1.85-3.34] | 2.62 [1.83-3.75] | 2.43 [1.49-3.95] |
| Social grade C2DE [ref ABC1] | 1.01 [0.77-1.32] | 1.16 [0.85-1.59] | 0.75 [0.46-1.23] |
| Minority ethnic group [ref white] | 1.81 [1.34-2.43] | 1.84 [1.30-2.60] | 1.69 [1.01-2.81] |
| Region in England [ref North] | | | |
| Midlands | 0.79 [0.55-1.13] | 0.63 [0.42-0.96] | 1.15 [0.63-2.09] |
| South | 0.99 [0.72-1.34] | 0.84 [0.58-1.20] | 1.26 [0.74-2.13] |
| Current vaping [ref not] | 4.07 [3.09-5.35] | 3.80 [2.74-5.27] | 4.76 [3.05-7.42] |
| Current cigarette smoking [ref not] | 3.74 [2.84-4.92] | 3.37 [2.43-4.67] | - |
| Smoking status [ref never regularly smoked] | | | |
| Former smoking | - | - | 3.14 [1.76-5.61] |
| Current smoking[5] | - | - | 8.07 [4.66-14.0] |

CI, confidence interval. OR, odds ratio.

[1] Defined as current use of pipes, cigars, cigarillos, waterpipes, bidis, snus, heated tobacco, or other smokeless tobacco.

[2] Also includes participants who reported (separately) that they did not currently smoke cigarettes but smoked some other form of tobacco.

[3] Any current use of pipes, cigars, cigarillos, waterpipes, or bidis.

[4] Any current use of snus, heated tobacco, or other smokeless tobacco.

[5] Current smoking includes participants who reported smoking cigarettes or other tobacco products.

**Table S7.** Unadjusted associations of any current use of different forms of non-cigarette tobacco with sociodemographic characteristics, vaping status, and smoking status

| | OR [95%CI] | | | | | | | |
|---|---|---|---|---|---|---|---|---|
| | Pipe | Cigar | Cigarillo | Waterpipe | Bidis | Snus | Heated tobacco | Other smokeless tobacco[1] |
| Age (years) [ref ≥65] | | | | | | | | |
| 18-29 | 1.12 [0.35-3.57] | 2.52 [1.29-4.93] | 3.06 [1.21-7.76] | 22.6 [5.35-95.0] | 3.50 [0.58-21.1] | 55.7 [7.62-407] | 7.84 [0.91-67.5] | 7.31 [1.48-36.2] |
| 30-64 | 1.04 [0.36-3.00] | 1.78 [1.00-3.18] | 1.93 [0.83-4.46] | 7.80 [1.85-32.8] | 0.46 [0.06-3.28] | 3.01 [0.36-25.3] | 4.17 [0.54-31.9] | 1.59 [0.32-7.91] |
| Men [ref women] | 3.14 [1.03-9.57] | 13.8 [5.38-35.5] | 3.57 [1.56-8.15] | 1.96 [1.15-3.36] | 0.77 [0.15-3.85] | 3.46 [1.76-6.82] | 1.13 [0.44-2.89] | 3.16 [1.18-8.46] |
| Social grade C2DE [ref ABC1] | 1.42 [0.62-3.27] | 0.88 [0.56-1.39] | 1.26 [0.67-2.38] | 0.93 [0.53-1.62] | 0.52 [0.06-4.22] | 0.50 [0.24-1.05] | 0.58 [0.18-1.85] | 1.63 [0.74-3.57] |
| Minority ethnic group [ref white] | 0.30 [0.06-1.35] | 1.58 [0.94-2.65] | 2.21 [1.10-4.42] | 7.92 [4.61-13.6] | 0.32 [0.04-2.72] | 1.58 [0.83-3.02] | 1.37 [0.42-4.46] | 2.10 [0.84-5.24] |
| Region in England [ref North] | | | | | | | | |
| Midlands | 0.24 [0.08-0.68] | 0.72 [0.40-1.30] | 0.55 [0.25-1.24] | 0.53 [0.25-1.12] | 0.60 [0.10-3.72] | 0.95 [0.43-2.09] | 2.00 [0.57-7.06] | 0.60 [0.20-1.82] |
| South | 0.23 [0.09-0.62] | 0.92 [0.56-1.53] | 0.53 [0.25-1.11] | 1.44 [0.80-2.60] | 1.13 [0.20-6.34] | 0.88 [0.46-1.70] | 1.97 [0.58-6.71] | 1.02 [0.41-2.54] |
| Current vaping [ref not] | 2.39 [0.94-6.10] | 4.76 [3.05-7.43] | 4.25 [2.21-8.17] | 5.86 [3.49-9.85] | 2.72 [0.58-12.7] | 4.73 [2.63-8.52] | 3.86 [1.51-9.87] | 5.16 [2.29-11.6] |
| Current cigarette smoking [ref not] | 3.90 [1.68-9.06] | 5.06 [3.27-7.81] | 13.0 [6.84-24.7] | 4.64 [2.74-7.88] | 4.14 [0.92-18.7] | - | - | - |
| Smoking status [ref never regularly smoked] | | | | | | | | |
| Former smoking | - | - | - | - | - | 2.36 [1.17-4.75] | 4.76 [0.86-26.4] | 6.57 [1.58-27.2] |
| Current smoking[2] | - | - | - | - | - | 3.60 [1.76-7.37] | 27.3 [6.09-123] | 28.6 [7.68-106] |
| Current nicotine pouch user [ref not] | - | - | - | - | - | 126 [65.0-246] | - | - |

CI, confidence interval. OR, odds ratio.

[1] Chewing tobacco, paan, gutka, snuff, and dip.

[2] Current smoking includes participants who reported smoking cigarettes or other tobacco products.
